# Quantification of arterial hemodynamics in steno-occlusive disease using time-resolved MRI-based angiography

**DOI:** 10.64898/2026.04.22.26350771

**Authors:** Quinten Deckers, Simone M. Uniken Venema, Kees Braun, A. Bart van der Zwan, Pieter T. Deckers, Jeroen C.W. Siero, Alex A. Bhogal

## Abstract

**Background:** Intracranial steno-occlusive disease (SOD) assessment benefits from hemodynamic imaging, but comprehensive evaluation often relies on contrast– or radiation-based techniques. Arterial spin labeling (ASL) provides a non-invasive alternative for quantifying tissue-level perfusion and cerebrovascular reactivity, yet does not capture upstream arterial flow dynamics. As a result, non-invasive assessment of macrovascular hemodynamics for SOD remains limited. This study evaluates whether quantitative 4D-MRA provides complementary arterial information beyond established ASL-derived metrics.

**Methods:** Twelve SOD patients (7 women; age 42.3±25.8 years) underwent multi-delay ASL and 4D-MRA before and after acetazolamide. Cerebrovascular reactivity (CVR), arterial transit time (ATT), macrovascular ATT (mATT), and labeled blood volume (LBV) were quantified. Associations and vasodilatory responses were assessed using linear mixed-effects models.

**Results:** At baseline, mATT correlated with ATT (β=0.66±0.08, p<0.001). Both decreased following acetazolamide (mATT: 1.07±0.03s to 1.01±0.03s, p=0.029; ATT: 1.63±0.07s to 1.40±0.07s, p<0.001). However, changes in mATT and ATT were not associated with CVR. In contrast, CVR was positively associated with ΔLBV (β=8.84, SE=2.43, p=0.01). Case analyses further demonstrated artery-level delayed inflow and vascular steal.

**Conclusion:** Quantitative 4D-MRA provides complementary macrovascular information to ASL in SOD. ΔLBV more consistently reflects cerebrovascular reactivity than transit-based metrics and is sensitive to artery-level delayed inflow and vascular steal.

*The local Medical Ethical Review Committee declared that the Medical Research Involving Human Subjects Act (WMO) did not apply (internal trial nr. 21-406)*.

## Introduction

In intracranial steno-occlusive disease (SOD) like moyamoya disease, hemodynamic information can support diagnosis, treatment planning, and follow-up^1,2^. For example, stenosis severity derived from structural MRI does not provide information on tissue-level cerebral blood flow (CBF) or autoregulatory compensation^3,4^. In our current clinical MRI protocols for SOD patients^5^, multi-delay arterial spin labeling (md-ASL) is used to quantify CBF, arterial transit time (ATT) and cerebrovascular reactivity (CVR) ^5–9^. When deciding on surgical revascularization, these data are considered alongside clinical parameters and digital subtraction angiography (DSA), which is done to assess stenotic severity and collateral pathways^10,11^. DSA is invasive and, while acceptable for initial diagnosis, unsuitable for serial monitoring^1,12^. For optimal patient selection, and to facilitate treatment monitoring and follow-up, a non-invasive hemodynamic workup would be ideal. This highlights a clinical need for radiation– and contrast-free MRI methods that deliver large-vessel hemodynamic information.

One candidate is ASL-based time-resolved magnetic resonance angiography (4D-MRA). 4D-MRA uses magnetically labelled blood to capture arterial flow dynamics across multiple inflow times. By fitting an appropriate signal model, macrovascular arterial transit time (mATT) can be quantified, which provides information that may reflect collateral routing and proximal flow impairment^13^. By introducing a vasoactive stimulus such as acetazolamide, the quantitative 4D-MRA protocol can inform on cerebrovascular reactivity (CVR) responses via changes in labelled blood volume (LBV) contained within the arterial tree. This follows from the fact that blood flow velocity increases such that 1) more spins should pass through the labelling slab, and 2) labelled spins will travel further down the vascular tree between the label and acquisition time, thus leading to a higher labelled blood volume. Under this formulation, the LBV may provide an arterial surrogate for the cerebrovascular reactivity (CVR) response at the level of the large intracranial vessels.

To evaluate the additional value of quantitative 4D-MRA–derived hemodynamic parameters, we assessed the concordance of mATT and LBV with perfusion-based reference metrics derived from multi-delay arterial spin labelling (ASL) in a clinical steno-occlusive disease population. We hypothesized that mATT would be shorter than ASL-derived ATT, reflecting proximal rather than tissue-level arrival, and that vasodilation would increase cerebral blood flow, shorten both transit time parameters, and increase LBV due to deeper arterial penetration of labelled blood. We additionally examined how regions affected by vascular steal might deviate from these patterns^14,15^.

## Materials and Methods

This single-center interventional study was performed at the University Medical Center Utrecht (UMCU; The Netherlands). The local Medical Ethical Review Committee declared that the Medical Research Involving Human Subjects Act (WMO) did not apply (trial nr. 21-406). All participants or their legal representative provided written informed consent to use their data. As this study is a proof-of-concept, no sample size calculation was performed.

### Study participants

Participants were included between July 2024 and March 2025 if they had confirmed SOD and a clinical indication for hemodynamic MRI as determined by the treating physician. All participants underwent MRI including ASL-based 4D-MRA and multi-delay ASL (md-ASL) before and after intravenous administration of acetazolamide within a single imaging session. Clinical indications included: evaluation of suitability for bypass surgery, assessment of cerebrovascular reactivity in the setting of new ischemic symptoms, or postoperative evaluation following neurosurgical intervention. Both paediatric and adult participants were eligible, irrespective of prior extracranial–intracranial bypass surgery. Only participants with complete pre– and post-acetazolamide ASL-based 4D-MRA and md-ASL datasets suitable for quantitative analysis were included. Participants were excluded if imaging was incomplete, degraded by severe motion artefacts, or if contraindications to MRI or acetazolamide were present (see “Participant Characteristics” in Results). Data from four participants were previously reported to introduce the mATT estimation model^13^; however, the present study evaluates mATT and LBV in relation to established ASL-derived parameters to assess sensitivity to pathological hemodynamic abnormalities.

### MRI protocol and data acquisition

All participants were scanned on a 3T scanner (Philips Healthcare, Best, the Netherlands) using a 32-channel receive coil (Invivo, Gainesville, USA). The clinical protocol^16^ contained a combination of standard structural imaging, md-ASL, and the newly added pseudo-continuous arterial spin labelling, combined with Keyhole and View-sharing scan (4D-PACK)^17^, which we refer to herein as 4D-MRA. The total scan duration was 45 minutes.

A dynamic pseudo-Continuous Arterial Spin Labeling (pCASL) sequence with a variable repetition time (vTR-ASL) and optimized background suppression was used for multi-delay ASL measurements. vTR-ASL enables flexible label durations and post-labeling delays, making it an efficient method for CBF and ATT assessment^13^. The ASL labeling plane was positioned manually to be perpendicular to the internal carotid and vertebral arteries. Arterial signal suppression was achieved using improved motion-sensitized driven-equilibrium with a TE of 16 ms and velocity encoding of 5cm/s in three directions^18^. The pCASL sequence contained a 3D GRASE readout, 10 label durations (300–2000 ms), and 10 post-labeling delays (200–3000 ms). Additional settings were turbo spin echo factor = 15, EPI factor = 15, SENSE factor = 2.5 × 1, half scan factor = 1 × 0.8 (right-left, feet-head direction), TR/TE range 1650–6150 ms/15 ms, flip angles = 90°/180°, voxel size = 4 × 4 × 8 mm³, field of view = 256 × 240 × 120 mm³, scan time = 2 min 54 s. The unique label duration and post-label delay combinations resulted in 10 saturation delays with a constant step size (500–5000 ms). A magnetization (M0) calibration image for CBF quantification was acquired with a 3000 ms TR. CBF and ATT quantification followed the Buxton kinetic model with nonlinear fitting^19^, using fixed parameters: labeling efficiency = 0.85, background suppression efficiency = 0.8, tissue T1 = 1.5 s, arterial blood T1 = 1.66 s, and tissue-blood partition coefficient = 0.9.

A vendor-specific (Philips) 4D ASL-based MRA sequence was used that incorporates ‘keyhole’ and ‘view-sharing’ techniques as part of the 4D-PACK implementation for scan acceleration^17^. Our parameters were as follows: 4D-MRA using a 3D GRE readout, 8 time-points at 100, 200, 400, 600, 800, 1200, 1600 and 2200 ms (label durations), 75% keyhole central size, turbo factor = 60, SENSE factor = 3 x 15 (right-left, feet-head direction), half scan (partial Fourier) factor = 0.8 x 0.8 (right-left, feet-head direction), TR/TE/flip angle = 6 ms/1.97 ms/11 degrees, acquired voxel size = 1 x 1.4 x 1.6 mm3, reconstructed voxel size = 0.6[x 0.6[x 0.8 mm^3^, field-of-view = 200 x 200 x 120 mm3, scan time = 6 min 16 s). Both the 4D-PACK and vTR-ASL sequences are part of Philips’ Advanced ASL Extension patch. After an intravenous injection of acetazolamide (20mg/kg with a maximum of 1g, dissolved in 30cc 0,9%NaCl, flow rate of 0.3 cc/s), a second multi-delay ASL was acquired, as well as a second 4D ASL-based MRA. The post-acetazolamide 4D-MRA and md-ASL scans were acquired 9±2 minutes and 16±2 minutes after injection, respectively.

### Image pre-processing and analysis

Multi-delay ASL data were processed using an in-house developed pipeline^20^ to generate pre-and post-acetazolamide CBF and ATT, using FSL BASIL for both the pre-acetazolamide and the post-acetazolamide ASL scan. CVR maps were calculated by subtracting pre-acetazolamide CBF maps from post-acetazolamide CBF maps (using the 2nd to the 5th post-label delay images). As such, ASL-CVR was defined as the absolute CBF change in ml/100 gr/min (= post-ACZ – baseline) and thus an increase in CBF from the baseline to the post-ACZ scan results in a positive ASL-CVR.

For an elaborate description of mATT fitting method, see Bhogal et al. (2025)^13^. Briefly, mATT was estimated using a piecewise saturation model, where an initial linear phase represents the arrival time of labelled spins, followed by a nonlinear saturation function describing signal increase. The model’s breakpoint indicates when the inflow signal surpasses the detection threshold, marking label-bolus arrival. More distal arterial voxels required longer label durations before reaching this threshold. Because 4D-MRA confines labelled spins to the arterial compartment, total labelled blood volume (LBV) and its acetazolamide-induced change (ΔLBV) were calculated as arterial markers of cerebrovascular reactivity. For each participant, all derived parameter maps (CBF, CVR, (Δ)ATT, (Δ)mATT, (Δ)LBV) were parcellated into 91 (sub)cortical regions using the native-transformed Harvard-Oxford atlas^21^. Regional averages were calculated for use in subsequent statistical analyses. The full preprocessing workflow is illustrated in Figure 1.

**Figure 1:**
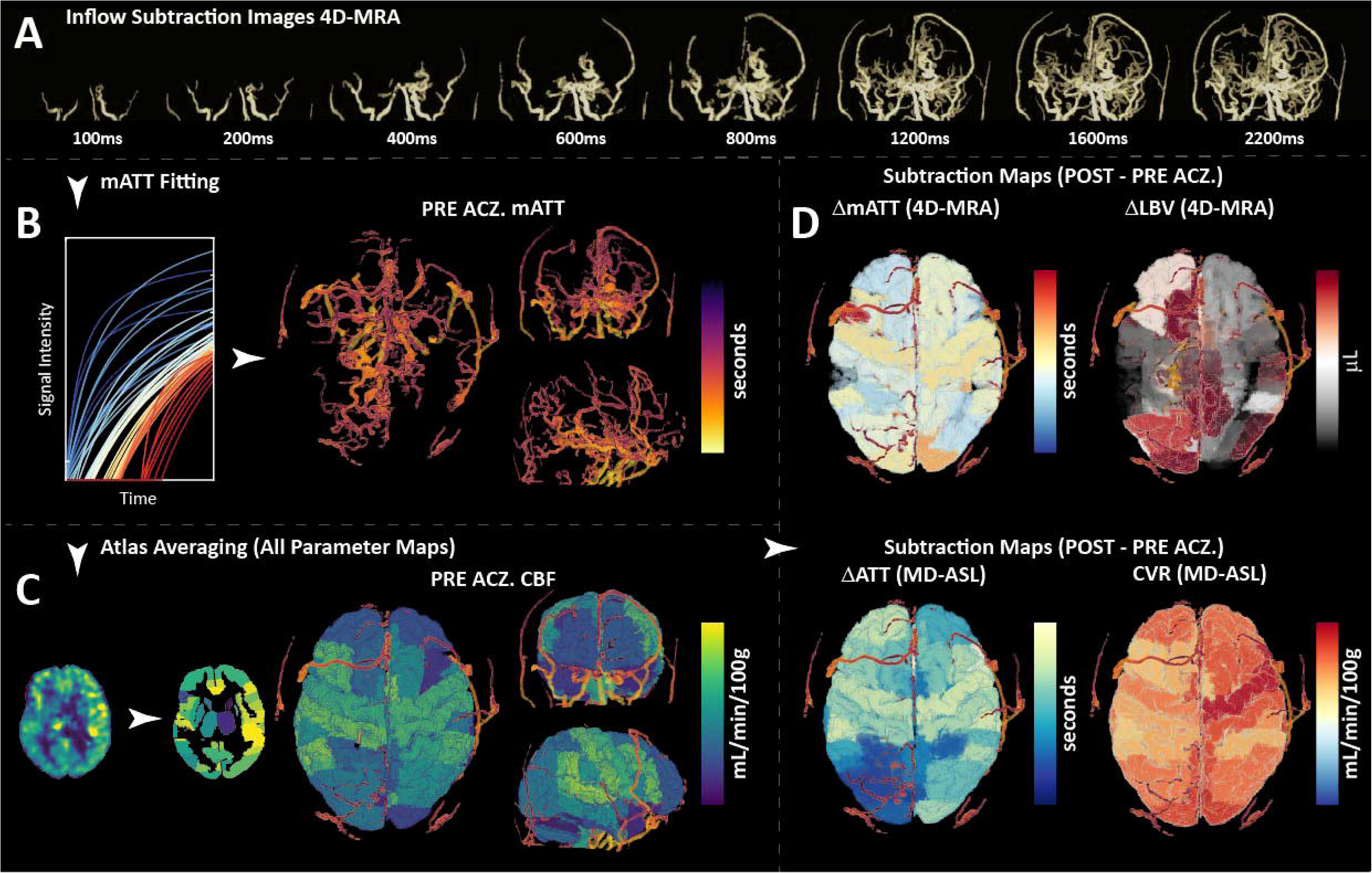
Preprocessing overview; A) Time-resolved inflow subtraction images from 4D-MRA illustrating progressive arterial filling; B) Pre-acetazolamide mATT maps generated by fitting a piecewise saturation model to arterial voxels; C) Atlas-based regional averaging of parameter maps, shown here for pre-acetazolamide CBF, and a 3D render of cortices of these maps with the macrovascular superimposed; D) Post–pre acetazolamide subtraction maps depicting vasodilatory changes in 4D-MRA–derived mATT and ΔLBV, alongside corresponding md-ASL derived ATT and CVR, with CVR defined as the absolute change in CBF, superimposed on a 3D render of the cortices of the parameter-based map. *4D-MRA = 4-dimensional magnetic resonance angiography, (m)ATT = (macrovascular) arterial transit time, CBF = cerebral blood flow, LBV = labelled blood volume, CVR = cerebrovascular reactivity, ACZ = acetazolamide.

### Statistical analysis

Linear mixed-effects (LME) models were used to assess associations between md-ASL and 4D-MRA parameters. We accounted for within-subject and within-region variability, with subjects and regions included as random effects. Age and sex were initially included as covariates to account for potential confounding. However, as these did not provide any meaningful contribution to the model, they were omitted from the final analyses to preserve statistical power and avoid overfitting. Model selection was guided by Akaike Information Criterion (AIC) to minimize bias in model selection (Table S1).

Three main analyses were performed in R (RStudio) using the lmer function^22^:

1. We assessed the correlation between mATT and ATT under the hypothesis that these correlate positively. We also expected a slight offset, as mATT measures upstream microvasculature compared to the tissue-level.
2. CVR was correlated with Δ(m)ATT to test the hypothesis that CVR varies inversely with arrival time as a result of increased perfusion after vasodilation.
3. CVR was correlated with ΔLBV to test the hypothesis that these metrics are positively correlated.

## Results

### Participant characteristics

Of the 18 eligible participants, four were excluded for poor 4D-MRA quality, one for poor md-ASL quality, and one for no consent to use their data (Figure 2). The final sample consisted of twelve participants (seven women; mean age=42.3±25.8 years) with SOD (Table 1). SOD was attributed to moyamoya disease (n=7), arterial occlusion (n=3), or atherosclerotic stenosis (n=1); in one participant, the diagnosis remained uncertain (possible moyamoya disease).

**Figure 2:**
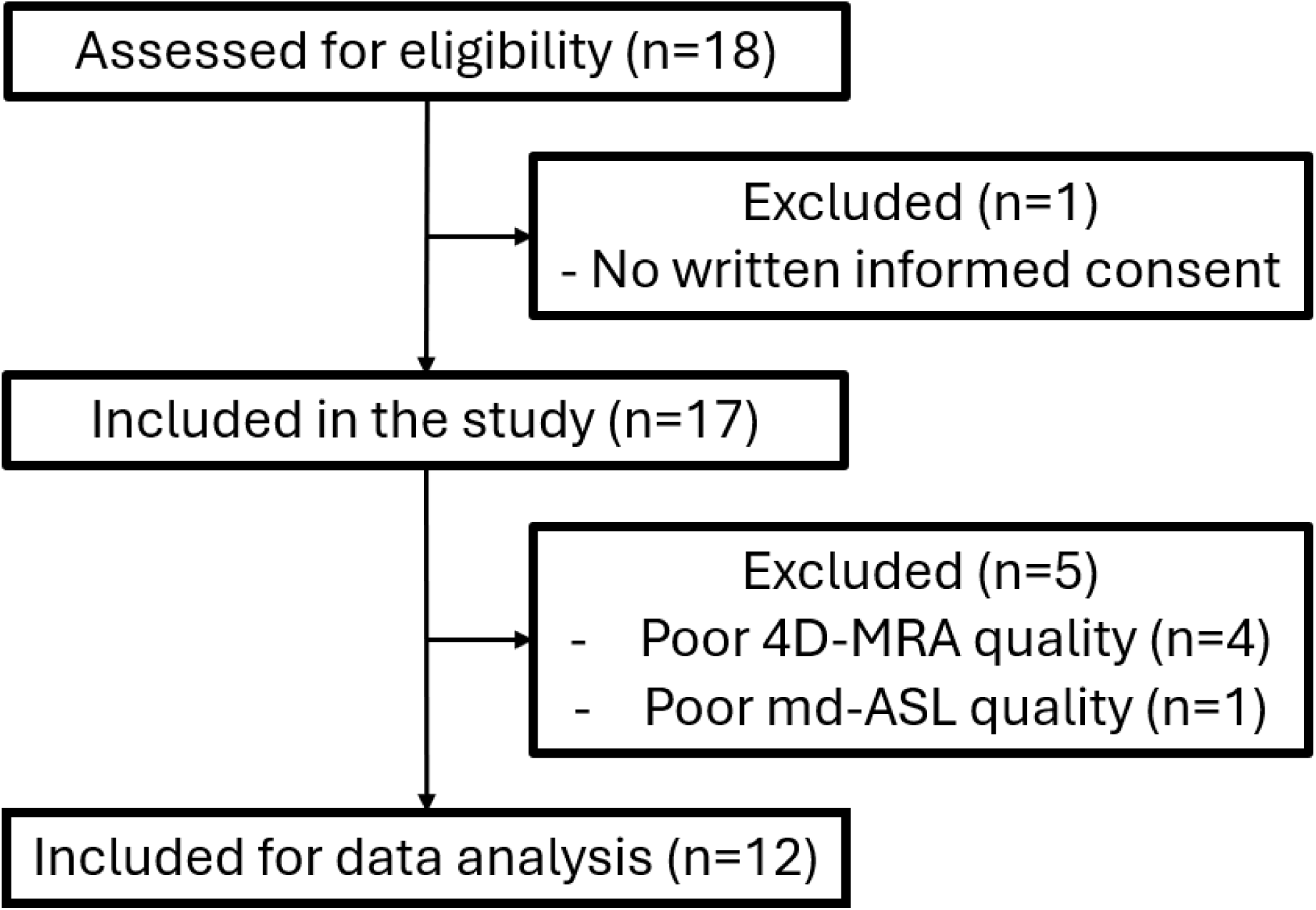
Flowchart shows inclusion for eligible participants with steno-occlusive disease. *Md-ASL = multi-delay arterial spin labeling, 4D-MRA = 4-dimensional magnetic resonance angiography.

**Table 1:**
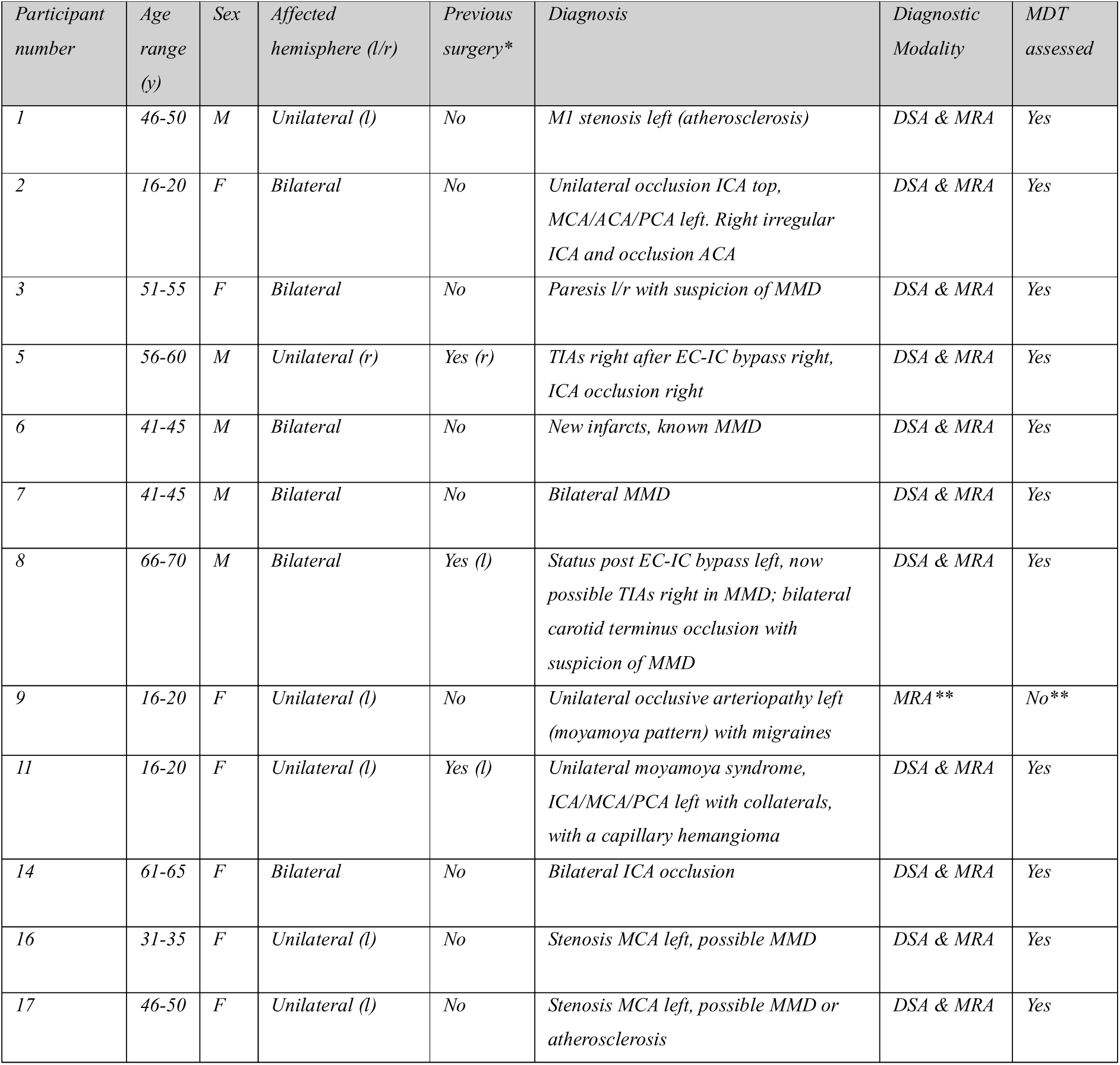
Additional participant information. *Surgery refers to the (l/r = left/right) hemisphere combined bypass. ** This pediatric participant only received an MRI and was discussed between a pediatric neurologist and vascular neurosurgeon. TIA = Transient Ischemic Attack, MMD = Moya-moya Disease, ICA = Internal Carotid Artery, MCA = Middle Cerebral Artery, ACA = Anterior Cerebral Artery, PCA = Posterior Cerebral Artery, EC-IC = Extracranial-intracranial, DSA = Digital subtraction angiography, MRA = Magnetic Resonance Angiography.

### Analysis 1: ATT versus mATT

ATT and mATT were positively associated both before (β=0.66, SE=0.08, 95% CI: 0.51 to 0.80, t=8.7, p<0.001), and after acetazolamide (β=0.77, SE=0.10, 95% CI: 0.58 to 0.96, t=7.8, p<0.001) (Figure 3A). The best-fitting model included random slopes for subjects and regions, reflecting intersubject heterogeneity and regional vascular differences. As mATT increased, pre– and post-acetazolamide ATT values converged. Significantly different intercepts (pre: β=0.87, SE=0.09, 95% CI: 0.68 to 1.05, p<0.001; post: β=0.56, SE=0.11, 95% CI: 0.35 to 0.77, p<0.001, difference: p<0.001) indicated a systematic offset between ATT and mATT.

**Figure 3:**
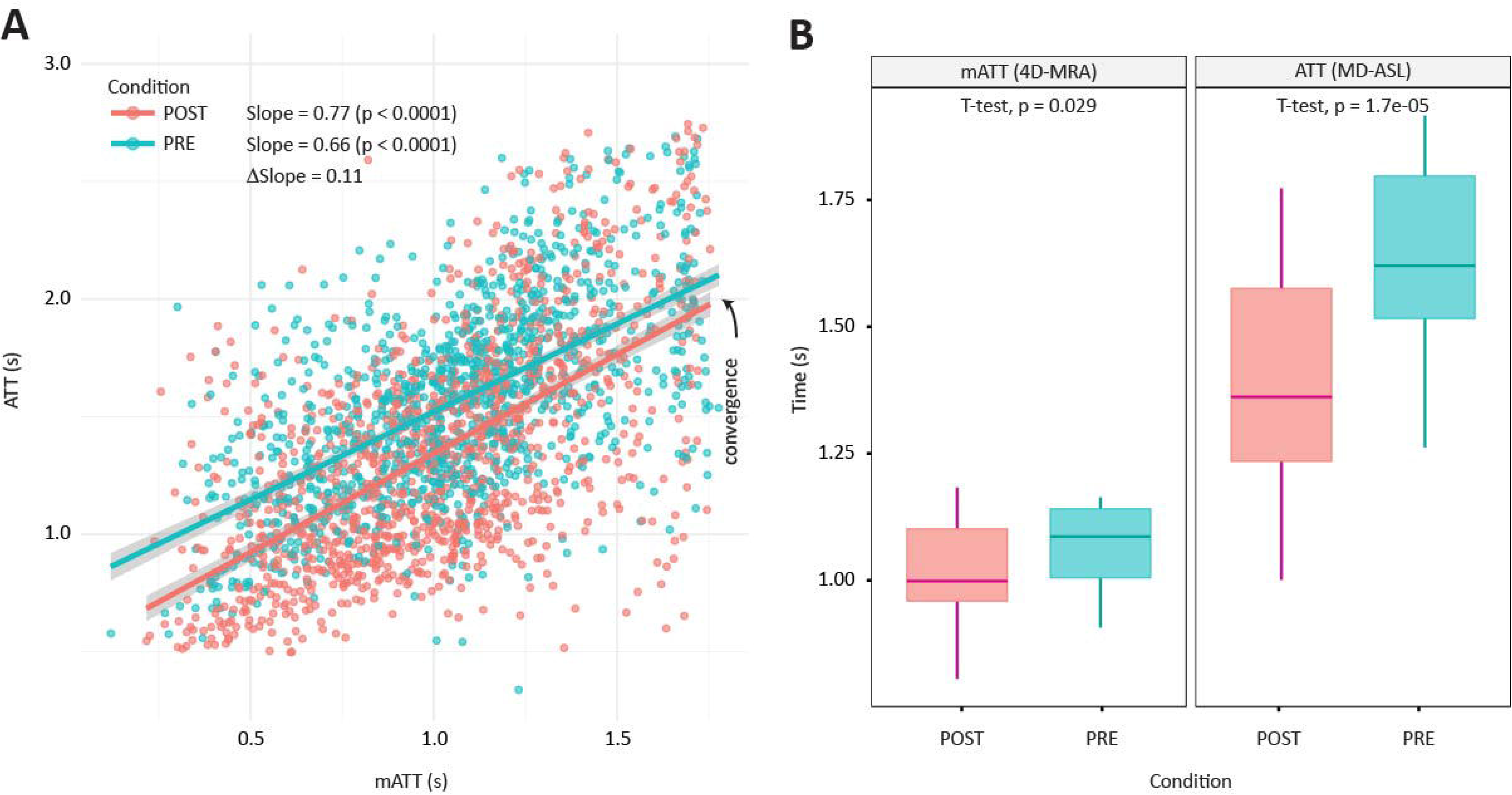
A) Linear mixed-effects model comparing ATT and mATT pre– (blue) and post-(red) acetazolamide. B) The effect of acetazolamide on both ATT and mATT reveal a shorter transit time in the post– condition (red) compared to the pre-acetazolamide condition (blue). As noted by the black arrow, the (m)ATT relationship converged at long transit times. *(m)ATT = (macrovascular) arterial transit time.

Furthermore, both mATT (pre: 1.07±0.03 sec; post: 1.01±0.03 sec; p=0.03) and ATT (pre: 1.63±0.07 sec; post: 1.40±0.07 sec; p<0.001) significantly decreased after acetazolamide, confirming the expected vasodilatory effect (Figure 3B). The reduction was larger for ATT than mATT.

### Analysis 2: CVR versus Δ(m)ATT

CVR showed no significant association with ΔATT (β=17.2, SE=8.4, 95%CI: –0.7 to 35.1, p=0.06) or ΔmATT (β=0.60, SE=0.44, 95%CI: –0.30 to 1.51, p=0.20). The ΔATT model explained 73% of the variance, though only 5.5% was attributed to ΔATT itself, indicating strong subject and regional variability. Participants with higher CVR generally showed lower or negative slopes (Figure 4A,B). On average, CVR was positive (95%CI: 13.4–14.9 ml/100gr/min), while ΔATT was negative (95%CI: –0.21 to –0.19s), consistent with increased flow and reduced transit time after vasodilation. For ΔmATT, the model explained 56% of the variance (R²<1%), with a similar pattern: positive CVR (14.2 ml/100gr/min, 95%CI: 13.4–14.9) and negative ΔmATT (95%CI: –0.085 sec to –0.060 sec). No significant relations were found when comparing CVR with baseline (m)ATT (ATT: p=0.27, mATT: p=0.91, Figure S2). When dividing unilateral cases (n=6) into affected and unaffected sides, no negative association was detected on either side (Figure S3).

**Figure 4:**
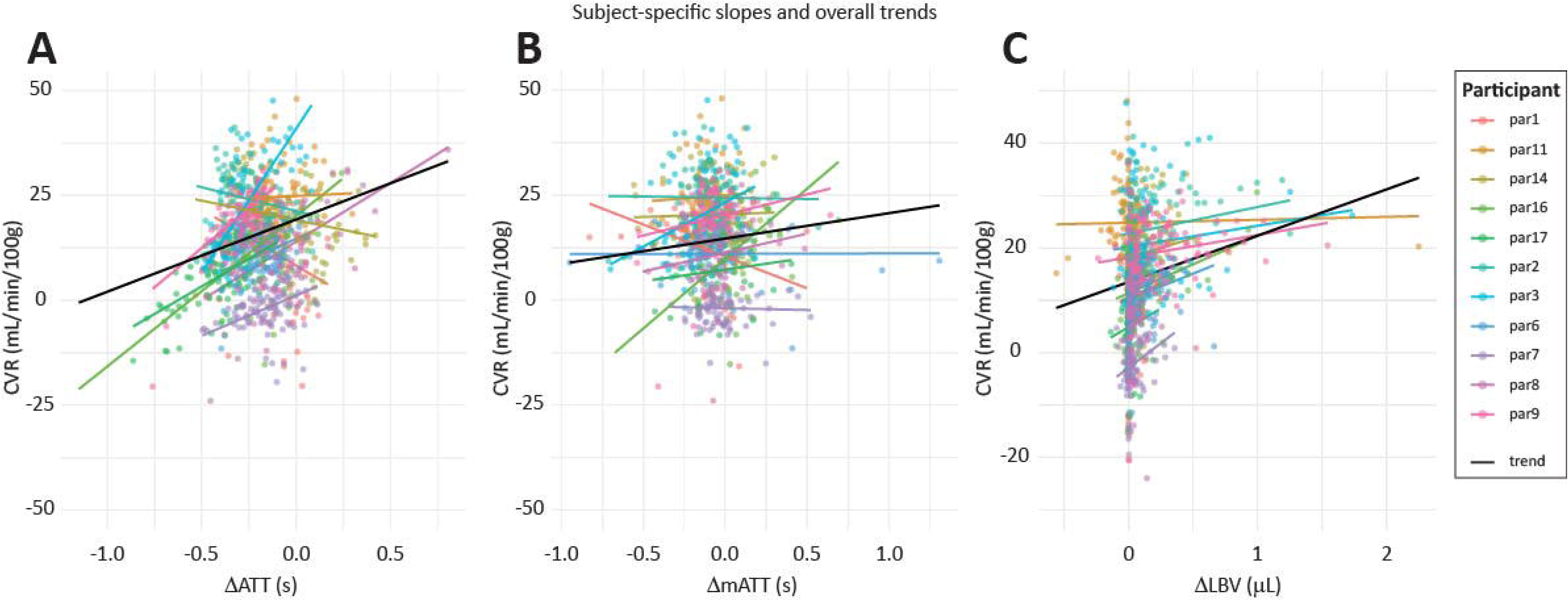
Subject-specific relationships from linear mixed-effects models between CVR and A) ΔATT from ASL, B) ΔmATT from 4D-MRA, and C) ΔLBV from 4D-MRA. Colored lines represent individual subject slopes with ROI-level data points; group-level trend is shown in black. *4D-MRA = 4-dimensional magnetic resonance angiography, Δ = post – pre acetazolamide, (m)ATT = (macrovascular) arterial transit time, LBV = labelled blood volume, CVR = cerebrovascular reactivity (= ΔCBF (cerebral blood flow)).

### Analysis 3: CVR versus ΔLBV

CVR and ΔLBV showed a positive correlation (β=8.84, SE=2.43, 95% CI: 4.1 to 13.6, p=0.01). Mean CVR was positive (14.2 ml/100gr/min, 95%CI: 13.4–14.9), and the average LBV increased by 0.15 mL (95%CI: 0.06–0.24) after acetazolamide. All but one subjects showed positive slopes (Figure 4C; sub11).

Some findings did not align with our original hypotheses. These are presented in short case reports below.

### Case 1

Negative CVR and ΔLBV (sub17): A woman in her 40s with a left M1 stenosis showed left-sided regions of low to negative CVR and ΔLBV following acetazolamide, indicating reduced perfusion and limited delivery of labelled blood after acetazolamide, compared to the pre-acetazolamide values. Correspondingly, vessels in the affected hemisphere exhibited prolonged mATT, whereas the contralateral hemisphere showed the expected mATT shortening with increased CVR and ΔLBV during vasodilation (Figure 5). This case illustrates how quantitative 4D-MRA can directly visualize arterial vascular steal, and thereby showing that CVR responds differently than stated in the hypothesis.

**Figure 5:**
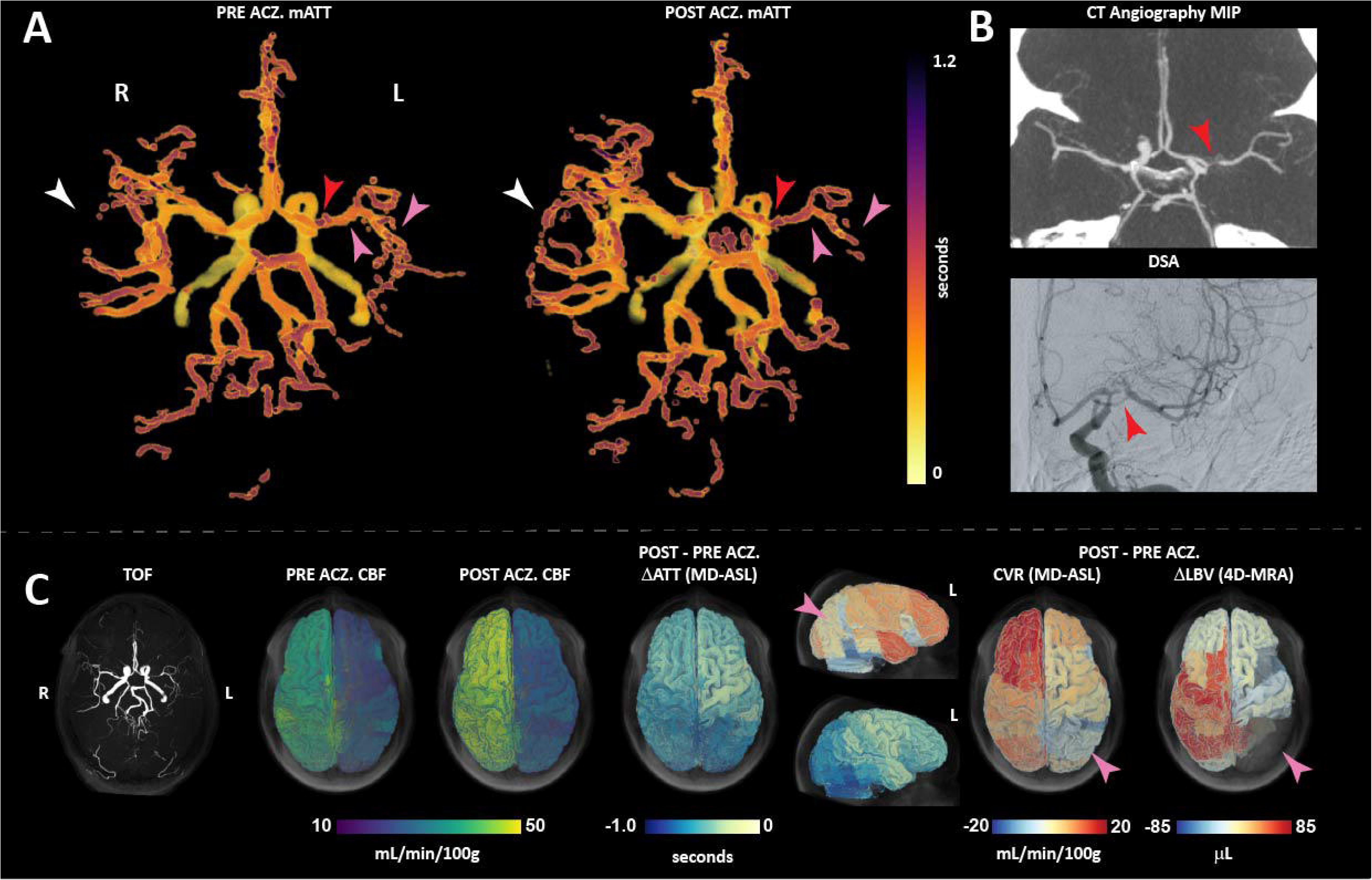
Vascular steal detected in a patient with a left M1 stenosis (red arrows); A) Pre-and post-acetazolamide mATT maps demonstrate left-sided prolongation of mATT with concomitant decreases in LBV, consistent with vascular steal (pink arrows). In the contralateral hemisphere, mATT shortens with increased LBV, as expected during vasodilation (white arrows); B) Maximum-intensity-projection CT angiography and digital subtraction angiography (DSA) of the circle of Willis confirm the left M1 stenosis (DSA: zoomed-in AP image with contrast in the left internal carotid artery (DSA: zoomed-in AP image with contrast in the left internal carotid artery); C) Time-of-flight MR angiography, CBF, ΔATT, CVR, and ΔLBV maps projected onto a cortical volume rendering using the SeeVieweR toolbox^21^ further illustrate vascular steal in the affected territory (pink arrows). *(m)ATT = (macrovascular) arterial transit time, CBF = cerebral blood flow, LBV = labelled blood volume, CVR = cerebrovascular reactivity, ACZ = acetazolamide, DSA = digital subtraction angiography, MIP = maximum intensity projection.

### Case 2

Elevated CBF with prolonged ATT (sub16): A woman in her 30s with a left M1 stenosis showed elevated regional CBF in the affected hemisphere despite prolonged ATT (Figure 6A).

**Figure 6:**
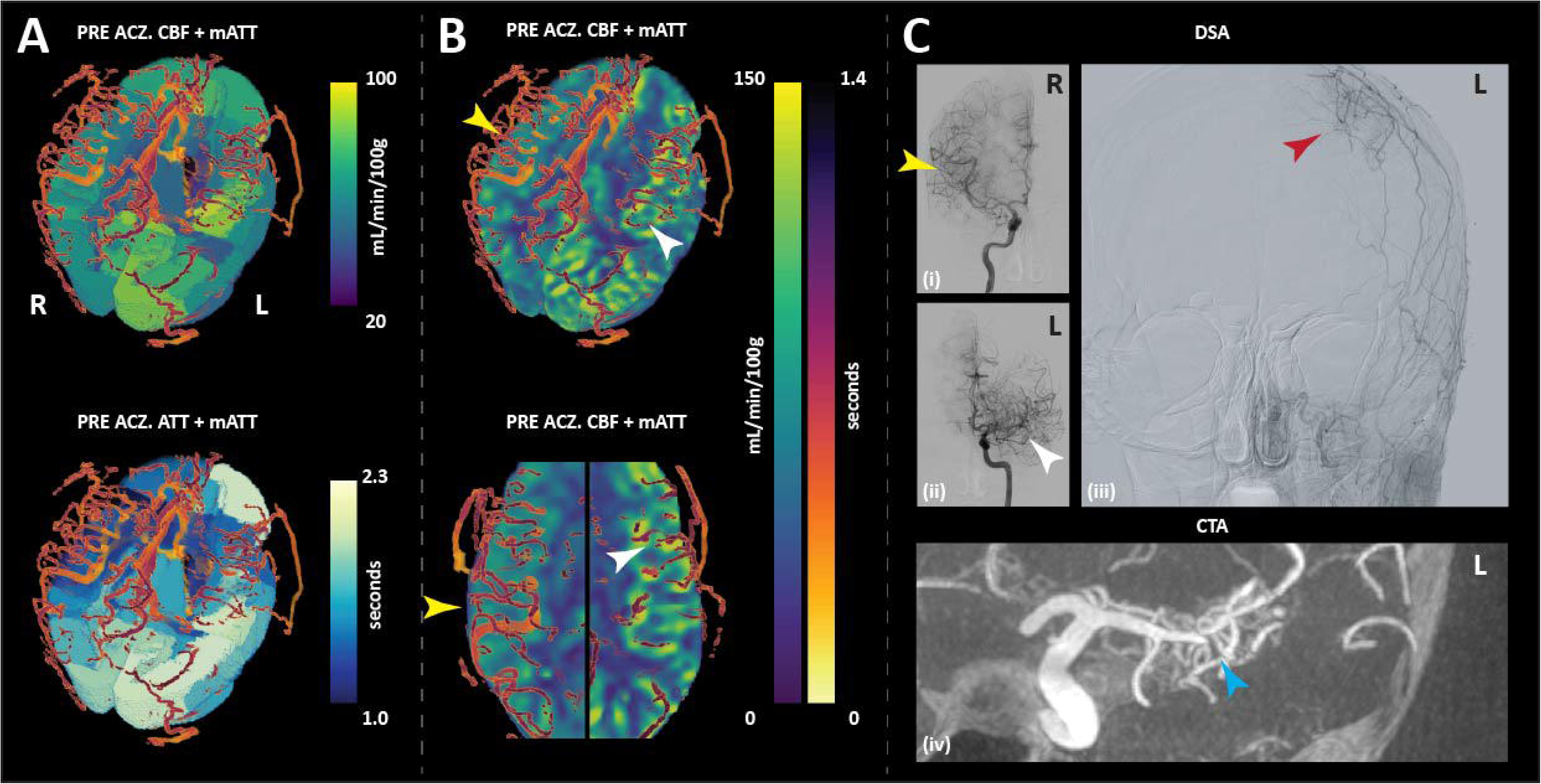
A) Volume-rendered regional-average CBF and ATT maps overlaid with mATT (created via the SeeVieweR Toolbox^21^) demonstrate similar spatial patterns, with regions of elevated CBF corresponding to prolonged ATT; B) Voxel-wise CBF maps show focal CBF hyperintensities that colocalize with large arteries exhibiting prolonged mATT, consistent with intravascular signal contamination (prolonged mATT: white arrows; unaffected CBF: yellow arrows); C) These artifacts likely arise from slow flow within collateral vessels associated with the M1 stenosis (blue arrow) and the external–internal carotid bypass (red arrow). *(m)ATT = (macrovascular) arterial transit time, CBF = cerebral blood flow, ACZ = acetazolamide, DSA = digital subtraction angiography, MIP = maximum intensity projection.

Voxel-wise CBF hyperintensities colocalized with arteries exhibiting prolonged mATT on 4D-MRA (Figure 6B), suggesting overestimation of regional CBF due to intravascular signal contamination, also known as arterial transit artifacts, despite the use of md-ASL. Slow arterial inflow was likely compensated by external–internal collateral pathways observed on the DSA (Figure 6C). This case illustrates how quantitative 4D-MRA provides complementary information that aids interpretation of complex hemodynamic patterns.

## Discussion

In this single-center study, we investigated the behavior of mATT and LBV derived from 4D-MRA during acetazolamide-induced vasodilation in participants with SOD. mATT demonstrated a strong correspondence with tissue-level ATT and shortened following acetazolamide administration, consistent with enhanced perfusion and preserved vasodilatory capacity. These findings support the physiological sensitivity of arterial-level transit metrics and align with prior reports^24,25^ describing reduced transit times after pharmacologic vasodilation.

Despite these concordant vasodilatory responses, we did not observe the anticipated inverse relationship between transit-time changes and CVR. This likely reflects the complex hemodynamic effects of acetazolamide: while vasodilation reduces transit time within individual proximal vessels, it simultaneously promotes deeper downstream penetration of labelled blood into distal arterial territories. The inclusion of distal arterial voxels with intrinsically longer transit times can bias region averaged mATT estimates (Figure S1). This effect is specific to mATT, which directly samples arterial signal, and cannot be observed for tissue-level ATT. Accordingly, pre– and post-acetazolamide mATT curves converged at longer transit times, indicating that regional averaging influences macrovascular transit estimates under vasodilatory challenge.

Further attenuation of the relationships between (m)ATT and CVR likely reflects pathological hemodynamic processes inherent to steno-occlusive disease. These include vascular steal phenomena, arterial transit artifacts affecting CVR estimation, and collateral recruitment, which is directly captured by macrovascular imaging but may obscure simple associations with tissue-level reactivity measures. In contrast, LBV remained positively associated with CVR, suggesting that LBV may represent a more robust physiological marker of enhanced perfusion and cerebrovascular reserve during vasodilatory challenge. Compared with transit-based metrics, LBV may be less sensitive to regional averaging effects and may better reflect the net capacity for flow augmentation.

Across participants, mATT values were systematically shorter than ATT, consistent with the expected temporal delay between macrovascular inflow and tissue-level arrival and in agreement with prior work^13^. Individual case examples further demonstrated concordant behavior of macrovascular (mATT and LBV) and tissue-level (CVR) metrics within defined regions of interest. Hemodynamically impaired regions exhibited negative CVR responses indicative of vascular steal, prolonged mATT, and reduced or absent ΔLBV following acetazolamide administration. In contrast, unaffected regions showed positive CVR, shortening of mATT, and increased LBV. This spatial concordance across modalities suggests that these parameters capture complementary manifestations of the same underlying hemodynamic state. The observed pattern likely reflects critically diminished inflow in compromised territories, such that labelled blood fails to adequately propagate into distal arterial segments within the imaging window. Notably, while LBV demonstrated robust detection of these flow deficits, the artery-specific nature of mATT provided enhanced spatial specificity for identifying pathological arterial segments. Collectively, these findings suggest that macrovascular 4D-MRA–derived metrics may provide greater specificity for detecting pathological arterial adaptations than tissue-level ATT alone, particularly in the presence of complex collateral flow patterns.

### Considerations

First, the modest sample size and the inherent heterogeneity associated with SOD meant that our statistical analysis were influenced by inter-individual variability. However, this also highlights that the 4D-MRA derived parameters are sensitive to individual differences, which is of note considering that treatment decisions in this population group are highly personalized. Nevertheless, future studies in larger, more homogeneous cohorts are warranted to evaluate ΔmATT and ΔLBV behaviour across specific cerebrovascular pathologies.

Second, the 4D-MRA acquisition preceded multi-delay ASL, which may have reduced the apparent vasodilatory response in ΔmATT if acetazolamide had not reached its maximal effect; this likely further increased variability but was unavoidable considering the clinical nature of our acquisition protocol. We note that recent scanner software innovations will facilitate further optimization of the 4D-MRA protocol. AI-based denoising combined with compressed sensing techniques allow to extend the acquisition window to increase inflow times, decrease overall scan time for easier clinical integration, or increase temporal sampling to yield more precise arrival-time estimates.

### Conclusion

4D-MRA–derived metrics enable artery-specific assessment of cerebrovascular hemodynamics in steno-occlusive disease. While the relationship between CVR and ΔmATT was influenced by collateral flow patterns, vascular steal, and transit-related effects, changes in LBV demonstrated a consistent positive association with CVR, indicating that ΔLBV may serve as a robust marker of arterial vasodilatory capacity. Together, these findings suggest that combined evaluation of ΔmATT and ΔLBV can provide additional insight into upstream arterial hemodynamics during vasodilatory challenge.

## Acknowledgements

We acknowledge the divisional support lead by Prof. Jeroen Hendrikse. We thank Jan Willem Dankbaar for clinical/neuro-radiological expertise and we are grateful to our nurses and MR technicians for their patient care and assistance in acquiring high-quality and reliable neuroimaging data.

## Data availability

Data generated or analyzed during the study are available from the corresponding author by request, as stated in the written informed consents of the participants.

## Abbreviations

SOD: Steno-Occlusive Disease
4D-MRA: 4 Dimensional Magnetic Resonance Angiography
(md/vTR/pC)-ASL: (multi-delay/variable repetition time/pseudo Continuous) Arterial Spin Labelling
(m)ATT: (macrovascular) Arterial Transit Time
LBV: Labelled Blood Volume
CBV: Cerebral Blood Volume
CVR: Cerebral Vascular Reactivity
CBF: Cerebral Blood Flow
DSA: Digital Subtraction Angiography
UMCU: University Medical Centre Utrecht
MNI: Montreal Neurological Institute (template)
LME: Linear Mixed-Effects
AIC: Aikaike Information Criterion
SE: Standard Error
TE: Echo Time
TR: Repetition Time

## Figure legends

**Figure S1:** Simulation illustrating why mean arterial transit time (mATT) does not necessarily show a negative association with cerebrovascular reactivity (CVR), despite evidence of increased penetration depth. (A) mATT angiogram of participant 3, highlighting an unaffected region selected for detailed inspection. (B) Arterial label propagation in this region before and after acetazolamide (ACZ) administration. With an identical acquisition time window, post-ACZ conditions show visibly increased penetration depth (white arrow), consistent with enhanced flow. (C) Conceptual illustration demonstrating how regional averaging attenuates the apparent mATT reduction. The blue curve represents pre-ACZ arterial label propagation, while the red curve represents post-ACZ propagation, characterized by faster and deeper penetration. At the penetration depth corresponding to pre-ACZ acquisition limit, the true mATT reduction is 0.80 s. However, when mATT values are averaged across the region up to the end of the acquisition window (≤ 2.2 s), the observed difference is reduced to only 0.22 s. This illustrates how spatial averaging can mask local transit-time shortening, explaining why mATT may fail to decrease despite increased CVR. Together with collateral recruitment, arterial transit artifacts, and vascular steal, these effects highlight the complexity of the relationship between CVR and mATT. *(m)ATT = (macrovascular) arterial transit time, CVR = cerebrovascular reactivity, LBV = labelled blood volume, ACZ = acetazolamide, AUC = area under the curve.

**Figure S2:** Subject-specific relationships derived from linear mixed-effects models between cerebrovascular reactivity (CVR) and baseline arterial transit metrics. A) CVR versus baseline macrovascular arterial transit time (mATT) measured with 4D-MRA (p = 0.91; β = 0.32 ± 2.8). A) CVR versus baseline arterial transit time (ATT) measured with arterial spin labeling (p = 0.27; β = −3.3 ± 2.9). Each data point represents a region-averaged value from an individual participant and is color-coded by participant. *(m)ATT = (macrovascular) arterial transit time, CVR = cerebrovascular reactivity, 4D-MRA = 4-dimensional MR-Angiography.

**Figure S3:** Subject-specific relationships derived from linear mixed-effects models in unilateral steno-occlusive disease. Analyses were restricted to unilateral cases, with regions classified as affected or unaffected. A–D) Associations between cerebrovascular reactivity (CVR) and transit metrics are shown for affected and unaffected regions: A) affected CVR versus ΔATT, B) unaffected CVR versus ΔATT, C) affected CVR versus ΔmATT, and D) unaffected CVR versus ΔmATT. In unaffected regions, a significant interaction between ΔmATT and ΔLBV was observed (p = 0.007; β = 0.011 ± 0.004), indicating that the relationship between ΔmATT and CVR becomes more positive with increasing labelled blood volume. This interaction was not present in affected regions and was not observed for CVR–ΔATT relationships. Also, a significant positive association between CVR and ΔATT was observed in unaffected regions (p = 0.002; β = 48.5 ± 7.8). *(m)ATT = (macrovascular) arterial transit time, CVR = cerebrovascular reactivity, LBV = labelled blood volume.

## Tables

**Table S1:**
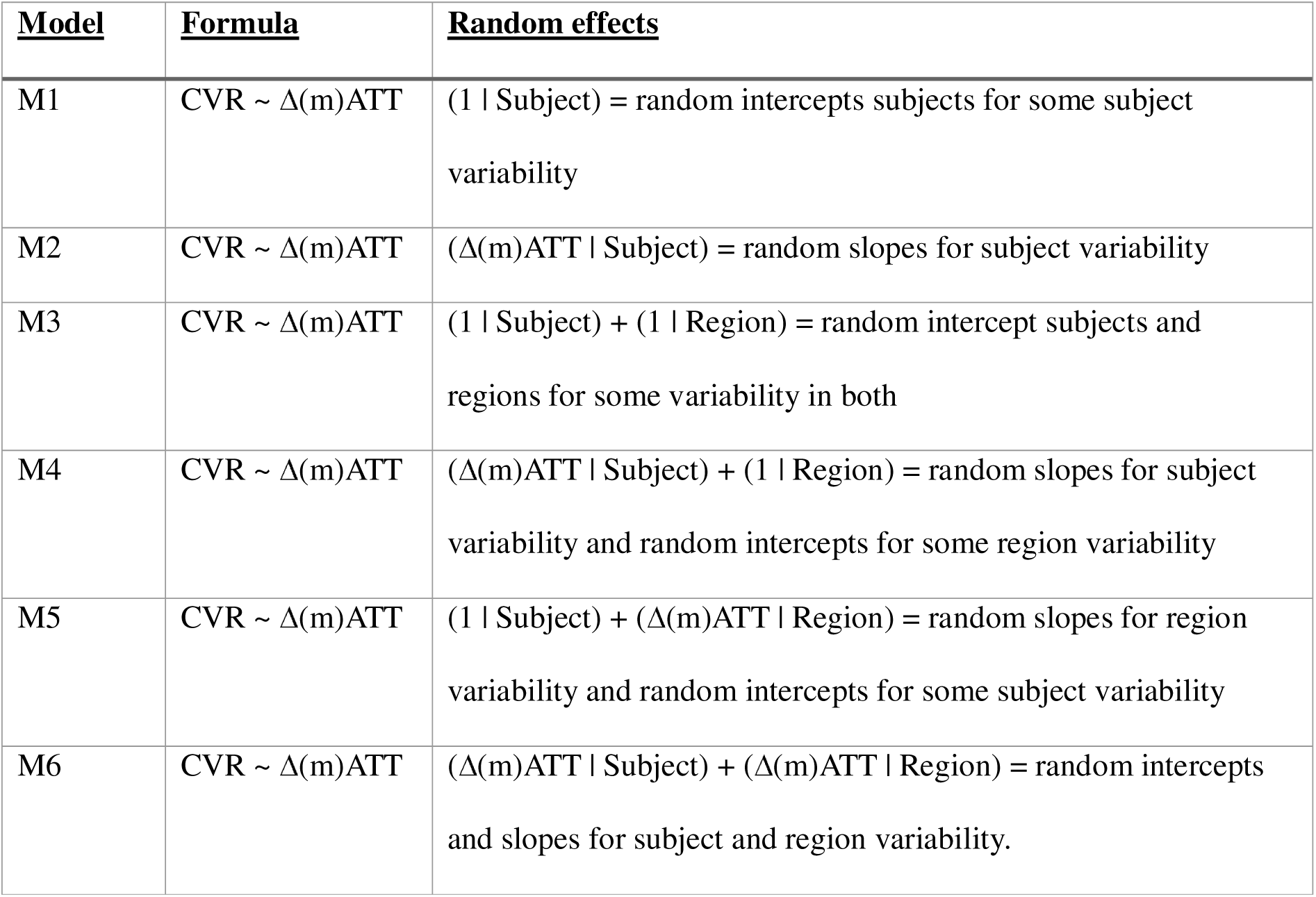
Additional model information. Before each linear mixed-effects model, the model with the lowest Akaike Information Criterium (AIC) was chosen. All three measurements that were performed showed that model M6 could explain the variance best. *(m)ATT = (macrovascular) arterial transit time, CVR = cerebrovascular reactivity.

