## Supplementary figures and images for "Quantification of arterial hemodynamics in steno-occlusive disease using time-resolved MRI-based angiography"

### Supplemental Figure 1

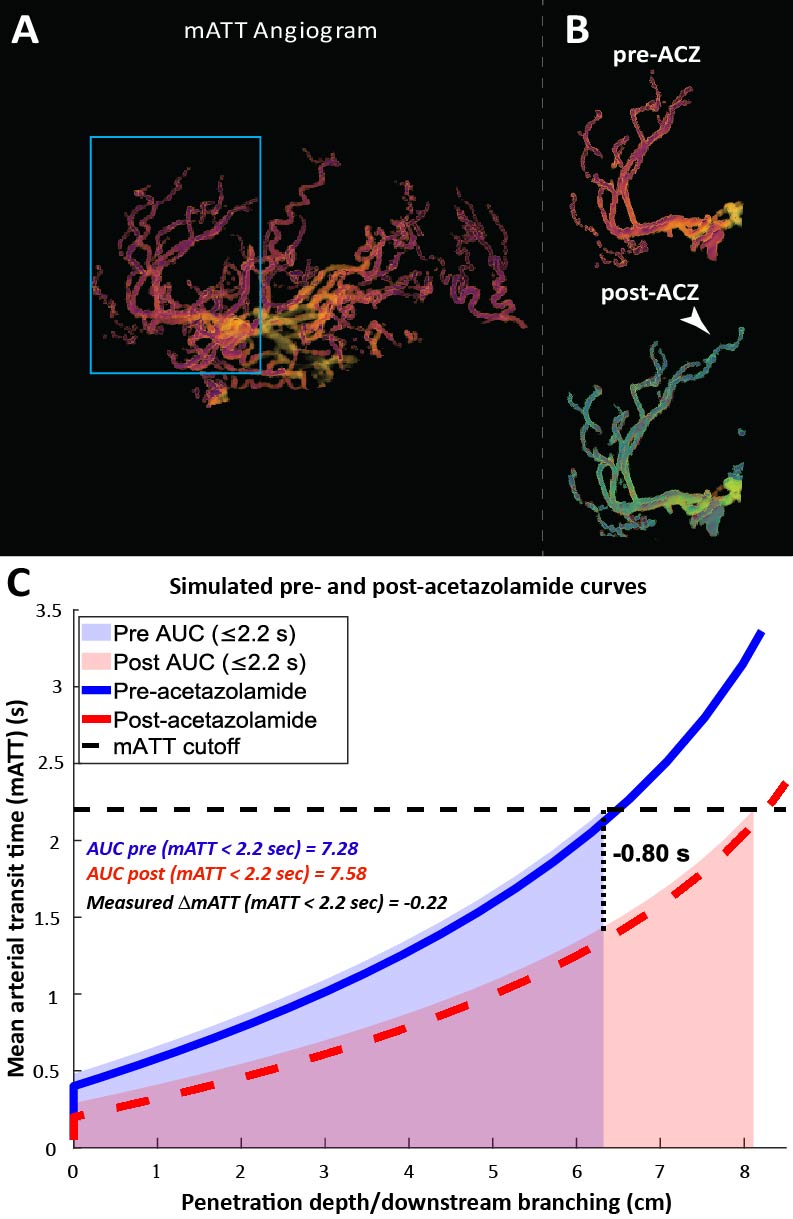

### Supplemental Figure 2

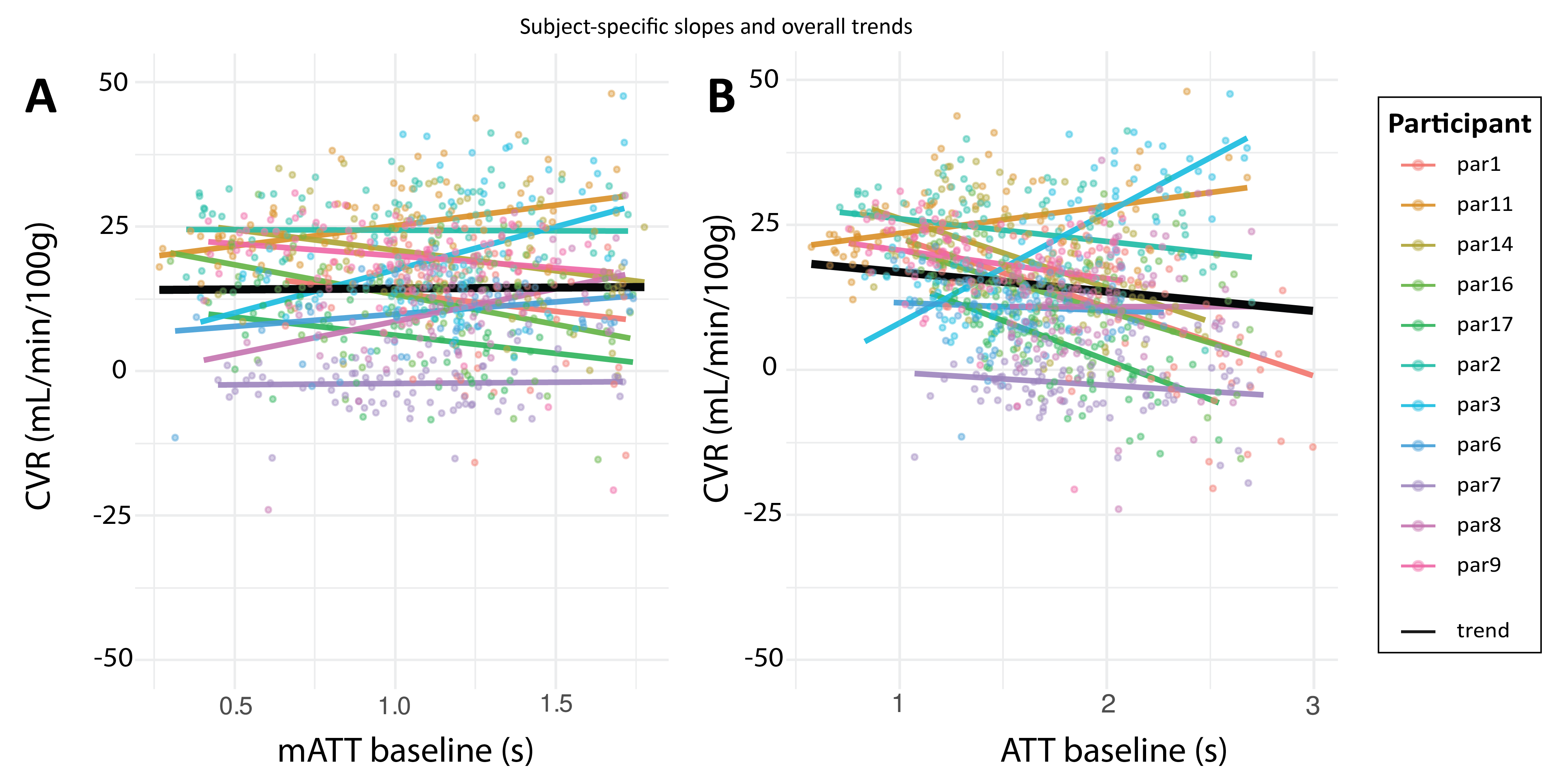

### Supplemental Figure 3

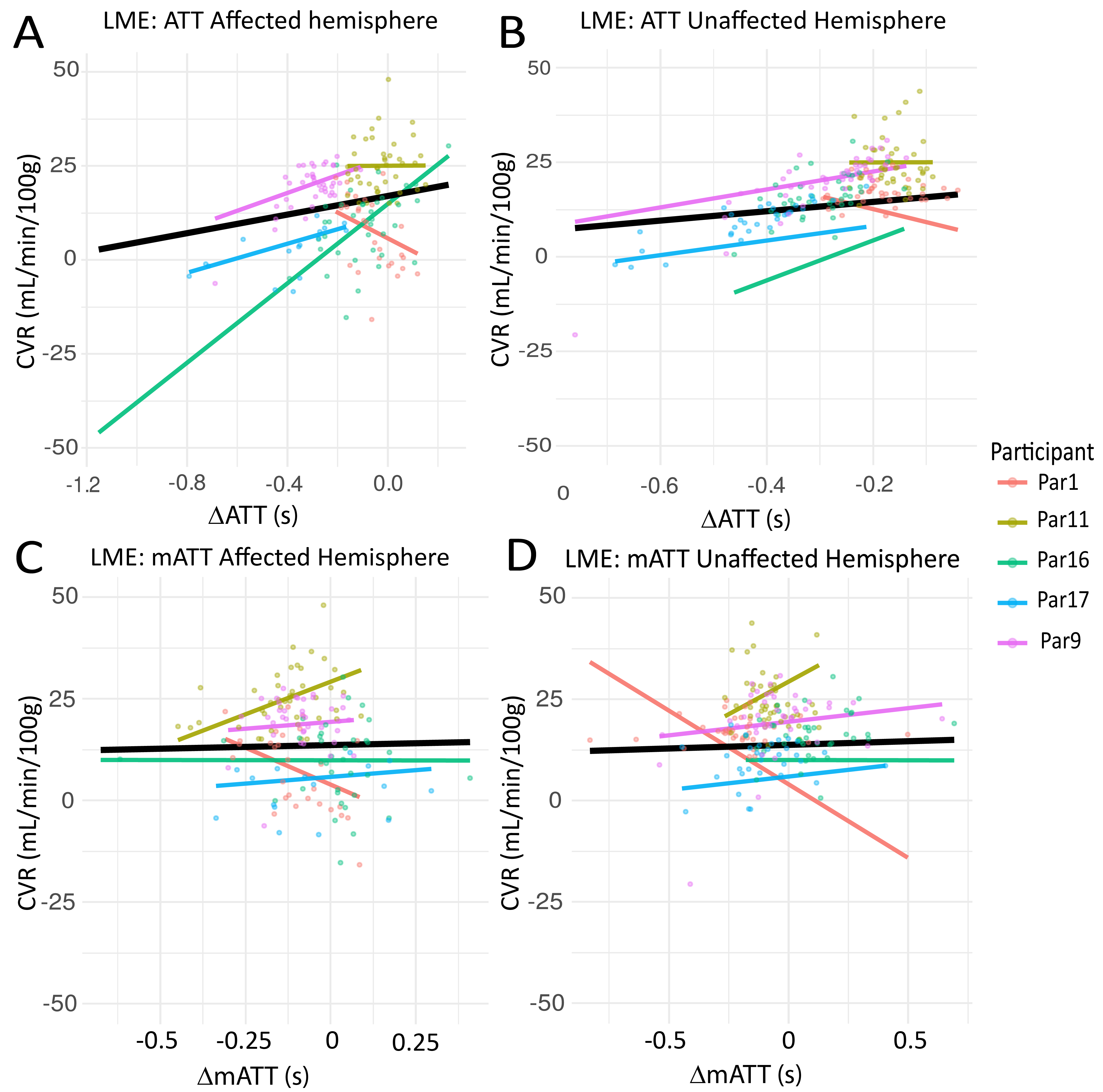
